# *GALC* variants affect galactosylceramidase enzymatic activity and risk of Parkinson’s disease

**DOI:** 10.1101/2022.04.30.22274239

**Authors:** Konstantin Senkevich, Cornelia E. Zorca, Aliza Dworkind, Uladzislau Rudakou, Emma Somerville, Eric Yu, Alexey Ermolaev, Daria Nikanorova, Jamil Ahmad, Jennifer A. Ruskey, Farnaz Asayesh, Dan Spiegelman, Stanley Fahn, Cheryl Waters, Oury Monchi, Yves Dauvilliers, Nicolas Dupré, Lior Greenbaum, Sharon Hassin-Baer, Francis P. Grenn, Ming Sum Ruby Chiang, S. Pablo Sardi, Benoît Vanderperre, Cornelis Blauwendraat, Jean-François Trempe, Edward A. Fon, Thomas M. Durcan, Roy N. Alcalay, Ziv Gan-Or

## Abstract

The association between glucocerebrosidase (GCase), encoded by *GBA*, and Parkinson’s disease highlights the role of the lysosome in Parkinson’s disease pathogenesis. Genome-wide association studies (GWAS) in Parkinson’s disease have revealed multiple associated loci, including the *GALC* locus on chromosome 14. *GALC* encodes the lysosomal enzyme galactosylceramidase (GalCase), which plays a pivotal role in the glycosphingolipid metabolism pathway. It is still unclear whether *GALC* is the gene driving the association in the chromosome 14 locus, and if so, by which mechanism.

We first aimed to examine whether variants in the *GALC* locus and across the genome are associated with GalCase activity. We performed a GWAS in two independent cohorts from a)Columbia University and b)the Parkinson’s Progression Markers Initiative study, followed by a meta-analysis with a total of 976 Parkinson’s disease patients and 478 controls with available data on GalCase activity. We further analyzed the effects of common *GALC* variants on expression and GalCase activity using genomic colocalization methods. Mendelian randomization was used to study whether GalCase activity may be causal in Parkinson’s disease. To study the role of rare *GALC* variants we analyzed sequencing data from 5,028 Parkinson’s disease patients and 5,422 controls. Additionally, we studied the functional impact of *GALC* knock-out on alpha-synuclein accumulation and on GCase activity in neuronal cell models and performed *in silico* structural analysis of common *GALC* variants associated with altered GalCase activity.

The top hit in Parkinson’s disease GWAS in the *GALC* locus, rs979812, is associated with increased GalCase activity (b=1.2; se=0.06; p=5.10E-95). No other variants outside the *GALC* locus were associated with GalCase activity. Colocalization analysis demonstrated that rs979812 was also associated with increased GalCase expression. Mendelian randomization suggested that increased GalCase activity may be causally associated with Parkinson’s disease (b=0.025, se=0.007, p=0.0008). We did not find an association between rare *GALC* variants and Parkinson’s disease. *GALC* knockout using CRISPR-Cas9 did not lead to alpha-synuclein accumulation, further supporting that increased rather than reduced GalCase levels may be associated with Parkinson’s disease. The structural analysis demonstrated that the common variant p.I562T may lead to improper maturation of GalCase affecting its activity.

Our results nominate *GALC* as the gene associated with Parkinson’s disease in this locus and suggest that the association of variants in the *GALC* locus may be driven by their effect of increasing GalCase expression and activity. Whether altering GalCase activity could be considered as a therapeutic target should be further studied.

## Introduction

The most recent genome-wide association study (GWAS) in Parkinson’s disease revealed multiple novel loci ^1^, most of them located in non-coding DNA ^2^. Since the top variants are in linkage disequilibrium with multiple other variants across multiple genes in each locus, in most cases it is unclear which variant and which gene is causally associated with the disease ^3^. The mechanisms behind the association of variants identified by GWASs are also mostly unknown. The effects of such variants can be mediated by changes in nearby gene expression, splicing, structural, biochemical or other functional properties of the translated protein, and they could be tissue-specific ^4,5^. Computational tools such as fine-mapping, genetic colocalization and Mendelian randomization, as well as functional studies, may help identify these variants and genes, and provide evidence for potential mechanisms ^6,7^.

There is mounting evidence about the role of lysosomal dysregulation in Parkinson’s disease ^8,9^. Variants in the lysosomal gene *GBA*, encoding the lysosomal enzyme glucocerebrosidase (GCase), are very common risk factors for Parkinson’s disease worldwide ^10-12^. GCase is an important enzyme in the glycosphingolipid metabolism pathway within the lysosome, and *GBA* risk variants in Parkinson’s disease are associated with reduced GCase activity ^13,14^. Other lysosomal genes and enzymes involved in this pathway have also been implicated in Parkinson’s disease, such as *SMPD1* ^15^, *ASAH1* ^16^ and *GLA* ^17^. Two recent GWAS meta-analyses have identified an association with Parkinson’s disease of a locus on chromosome 14 encompassing *GALC*, but also additional genes ^1,18^. *GALC* encodes galactosylceramidase (GalCase), an important enzyme in the glycosphingolipid metabolism pathway responsible for the degradation of galactosylceramides and galactosylsphingosines ^19^. However, it is not clear whether *GALC* is the gene associated with Parkinson’s disease in the locus on chromosome 14, and whether this association might be related to its activity in the glycosphingolipid metabolism pathway. There are currently several drugs in clinical trials targeting *GBA* and the glycosphingolipid metabolism pathway ^20^, and it is important to identify other druggable targets in this pathway for future development.

In this study, we aimed to examine whether common *GALC* variants that are associated with Parkinson’s disease, as well as other variants in this locus and across the genome, affect GalCase activity, and whether GalCase activity itself may be associated with Parkinson’s disease. First, we performed a GWAS on GalCase activity in two independent cohorts, followed by a meta-analysis with a total of 1,123 Parkinson’s disease patients and 576 controls. We further performed colocalization analyses of the *GALC* locus with its expression, as well as with GalCase activity. We used Mendelian randomization to examine whether GalCase activity may be causally associated with Parkinson’s disease. To study whether rare *GALC* variants may also be associated with Parkinson’s disease, we analyzed sequencing data from 5,028 Parkinson’s disease patients and 5,422 controls. Additionally, we studied the functional impact of *GALC* knock-out on alpha-synuclein accumulation and on GCase activity in human neuronal cell models and performed *in silico* structural analysis of GalCase variants.

## Materials and methods

### Study population

GWAS was performed in two cohorts with available data on enzymatic activity. The Columbia University cohort (New York) and the Parkinson’s Progression Markers Initiative (PPMI) cohort (detailed in Table 1). Both cohorts have been previously described in detail ^14,21,22^. Rare variants analysis was performed in three cohorts sequenced at McGill university from Columbia University, McGill University (Quebec, Canada and Montpellier, France) and Sheba Medical Center (Table 1). In addition, rare variants analysis was performed in the Accelerating Medicines Partnership – Parkinson Disease (AMP-PD) initiative cohorts (https://amp-pd.org/). The AMP-PD analysis included 2,607 Parkinson’s disease patients and 3,797 controls from the BioFIND study, Harvard Biomarkers Study (HBS), National Institute of Neurological Disorders and Stroke (NINDS) Parkinson’s disease Biomarkers Program (PDBP), PPMI, and the NINDS Study of Isradipine as a Disease Modifying Agent in Subjects With Early Parkinson Disease, Phase 3 (STEADY-PD3) and the National Institute on Aging (NIA) International Lewy Body Dementia Genetics Consortium Genome Sequencing in Lewy body dementia case-control cohort (Table 1). All Parkinson’s disease patients were diagnosed according to either the UK Brain bank criteria ^23^ or the Movement Disorders Society (MDS) criteria ^24^. All participants signed informed consent forms in their respective cohorts and the study protocol has been approved by the institutional review boards.

**Table 1.** Demographic data of the cohorts to study GalCase activity and rare variants in *GALC*.

| Cohort | Parkinson's disease (males %) | Controls (males %) | Parkinson's disease age, mean (SD in years) | Controls age, mean (SD in years) |
| --- | --- | --- | --- | --- |
| **Demographic data of the cohorts to study GalCase activity** |  |  |  |  |
| Columbia | 649(65.3%) | 337 (39%) | 64.20 (10.69) | 65.58 (11.07) |
| PPMI | 327 (67%) | 141 (68%) | 62.01 (9.41) | 61.32 (10.88) |
| **Demographic data of the cohorts to study rare variants in *GALC*** |  |  |  |  |
| Columbia | 954 (65%) | 506 (36%) | 65.73 (10.54) | 64.32 (10.06) |
| Sheba | 686 (61%) | 549 (69%) | 63.11 (12.29) | 33.94 (8.18) |
| McGill | 781 (62%) | 570 (44%) | 59.09 (10.77) | 55.22 (12.49) |
| AMP-PD | 2341 (62%) | 3486 (47%) | 64.66 (9.73) | 68.39 (13.43) |

SD – standard deviation; PPMI – Parkinson's Progression Markers Initiative; AMP-PD – Accelerating Medicines Partnership – Parkinson Disease

### Enzymatic activity

GalCase enzymatic activity was measured from dried blood spots (DBS) ^25^. In the Columbia cohort, DBS have been collected and analyzed as previously described ^14,26^. In brief, GalCase activity was measured in all available samples at Sanofi laboratories using liquid chromatography-tandem mass spectrometry (LC-MS/MS) from DBS, incorporated in a multiplex assay. DBS were incubated with a reaction cocktail containing substrates for lysosomal enzymes and buffer to maintain the reaction pH. The calculation of enzyme activity was carried out on the assumption that the amount of the obtained product after incubation with the substrate is directly proportional to the activity of lysosomal enzymes in a DBS ^26^. In the PPMI cohort, GalCase activity was measured using a similar method, after thawing frozen blood collected in EDTA tubes (kept at -80C) as previously described^21^. In brief, frozen whole blood was slowly thawed on watered ice within 45-60 minutes. Once thawed, DBS from frozen blood was prepared and stored in individual sealed plastic bags with a desiccant at -80C until analysis. Most of the participants from the PPMI cohort have provided blood samples for three consecutive years. The mean activity across all time points was calculated for the PPMI cohort and was used for further analyses as previously described ^21^. Outliers with activity > ± 3 z-scores were excluded from the analysis. We applied logistic regression models in R to compare activity in cases and controls.

### Genome wide association analysis

Genotyping was performed using the OmniExpress GWAS array according to the manufacturer’s instructions (Illumina Inc.). Quality control was performed on both individual and variant-level data as previously described (https://github.com/neurogenetics/GWAS-pipeline). In brief, we used the unimputed data to filter out heterozygosity outliers with F-statistics cut-off LJ>LJ± 0.15, samples with low call rate (<95%), and samples with a mismatch between the reported and genetically identified sex. On the variant level data, we excluded variants with high missingness and variants deviating from Hardy-Weinberg Equilibrium (P-value<1E-5). We performed imputation using the Michigan imputation server with the Haplotype Reference Consortium reference panel r1.1 2016 under default settings ^27^. In the GWAS analysis, we only included hard-call variants (R^2^>0.8) with minor allele frequency (MAF) > 0.01. GWAS was performed using logistic regression in plink version 1.9 adjusting for sex, age, disease status and 5 principal components ^28^. In the PPMI cohort, we also adjusted for white blood cell count as suggested previously ^21^. Conditional and joint analyses (COJO) were performed to identify independent single nucleotide polymorphisms (SNPs) in the GWASs after adjusting for the top hits ^29^. GWAS meta-analysis between the Columbia and PPMI cohorts was conducted using the METAL package in R ^30^. Mirror Manhattan plots were created with the Hudson R package (https://github.com/anastasia-lucas/hudson).

### Colocalization analysis

Genomic colocalization analysis allows for fine-mapping of genetic loci and provides an estimation of the overlap between risk variants and quantitative trait locus (QTL) variants. In other words, colocalization allows for determining whether the same variants that affect risk also affect traits such as expression of the nearby genes. We used the LocusCompareR R package to plot GWAS-eQTL colocalization events (https://github.com/boxiangliu/locuscomparer). Colocalization analysis was performed using the coloc R package (https://chr1swallace.github.io/coloc/index.html) ^31^. To perform colocalization we extracted variants from the region +/-500 kb around *GALC*. As a reference, to use the largest dataset available, we utilized the recent full Parkinson’s disease GWAS summary statistics including data from 23andMe^1^ and GalCase activity GWAS summary statistics derived from previous analysis. As a reference for eQTL we used a recent large-scale brain eQTL meta-analysis ^32^. Colocalization analysis considers five hypotheses: H0 - no association with Parkinson’s disease or QTL in the region; H1 - association with Parkinson’s disease only; H2 - association with QTL only; H3 - both Parkinson’s disease and QTL are associated with the studied region but have different and independent single associated variants; H4 - both Parkinson’s disease and QTL are associated and share the same single associated variant. We considered the colocalization analysis as significant if the posterior probability of colocalization in H4 (PPH4) was > 0.8 ^31^.

### Mendelian randomization

Mendelian randomization (MR) is a method that allows for testing potential causality between different traits using genetic data. We performed MR to study the potential causal relationship between the levels of GalCase activity and Parkinson’s disease. As exposure, we used the GWAS summary statistics that we generated for GalCase activity in the Columbia cohort, the PPMI cohort and the meta-analyzed data. Parkinson’s disease risk GWAS summary statistics were used as an outcome ^1^. Genetic instruments for the exposure were constructed using only GWAS significant SNPs (p-value < 5×10^−8^). We used a clumping window of 10,000 kb and the r2 threshold was set to 0.02.

To exclude instruments that explain more variance in the outcome (PD risk) than in exposure (GalCase activity), we applied Steiger filtering ^33^. The two-sample MR R package was used to perform MR ^33,34^. The MR-Egger method was used to account for directional pleiotropy and to estimate the true causal effect ^35^. We used the inverse-variance weighted (IVW) method to aggregate and meta-analyze estimates from individual Wald ratios for each SNP ^36^. To identify invalid instruments due to the horizontal pleiotropy, Cochran’s Q statistic implemented in IVW and MR-Egger methods was used. The MR pleiotropy residual sum and outlier (MR-PRESSO) global test was also used to detect horizontal pleiotropy ^37^.

### Gene burden analysis

We performed full sequencing of *GALC* in the cohorts from Columbia University, McGill University and Sheba Medical Center, using targeted next-generation sequencing with Molecular Inversion Probes (MIPs), as previously described ^38^. The quality control was performed as previously described ^38^, with minimal coverage of 30X for variant calls. The full protocol is available at https://github.com/gan-orlab/MIP_protocol and the code is available at https://github.com/gan-orlab/MIPVar/.

Whole-genome sequencing data was available through the AMP-PD portal ^39^. Quality control on whole-genome sequencing performed by AMP-PD on individual and variant levels was previously described (https://amp-pd.org/whole-genome-data). We have included only individuals with European ancestry from AMP-PD cohorts as there were not enough participants of other ethnicities to perform a meaningful analysis.

Genotype data from the McGill cohorts were annotated with hg19, and *GALC* coordinates were chr14:88,399,358-88,459,615; genotype data from AMP-PD were annotated with hg38, and coordinates for extraction were chr14:87,933,014-87,993,182. To meta-analyze these cohorts we used the LiftOver package to convert all genome positions to hg19 (https://genome.sph.umich.edu/wiki/LiftOver). To study the burden of rare variants (minor allele frequency < 0.01), we used the optimized sequence kernel association test (SKAT-O) and metaSKAT R packages ^40,41^. SKAT-O analysis of the *GALC* gene was performed separately for different types of variants: 1) all rare variants 2) all variants with a Combined Annotation Dependent Depletion (CADD) score ≥ 20 (representing 1% of the top deleterious variants) 3) all non-synonymous variants and 4) all functional variants including non-synonymous and loss-of-function variants (stop gain/loss, frameshift, and splicing variants located within two base pairs of exon-intron junctions).

### In silico structural analysis

The atomic coordinates of the full-length human GalCase protein were retrieved from the AlphaFold server and compared with the structure of mouse GalCase bound to 4-nitrophenyl beta-D-galactopyranoside (PDB 4CCC). The figure was generated using PyMol v.2.4.0.

### Generation and differentiation of human induced pluripotent stem cell (iPSC) lines

CRISPR-Cas9 genome editing was used to individually ablate the *GALC* or the *GBA* loci in AIW002-02 iPSCs ^42^. Quality control of the parental line was carried out as previously described ^42^. A pair of guide RNAs expressed together with Cas9 nuclease from PX459 (Addgene #48139) by transient transfection with Lipofectamine Stem (ThermoFisher Scientific, STEM00001) was used to target each locus in iPSCs: GGCTGGGAAAAGGTTTCGAC and GTCCAAATCATGGTAACGCT for the *GALC* locus; TAAAAGCTTCGGCTACAGCT and GCTATGAGAGTACACGCAGT for the *GBA* locus. Following puromycin selection, colonies were picked and screened by PCR and sequencing. The sequences of the screening primers were: TTGGTAAGGGTCTTGGAGAGA and AAACCCAGCTCAGAGGAAGG for the *GALC* locus; TTTTGGCTCATTCCAACCTC and TTGAGAGCAGCAGCATCTGT for the *GBA* locus. Both knockout lines were screened for pluripotency by immunofluorescence and for genomic integrity using the hPSC Genetic Analysis Kit (Stemcell Technologies, 07550) as described ^42^. The wild type and knockout iPSC lines were subsequently transduced with lentiviruses to generate lines that had the capacity to differentiate into Ngn2-induced neurons based on a protocol adapted from Zhang *et al*. and Meijer *et al*. and characterized by immunofluorescence for neuronal markers (data not shown) ^43,44^. *SNCA* triplication, Isogenic control (Isog Ctl) and *SNCA* KO NPCs were differentiated by the monolayer method from iPSCs ^45^.

### GCase activity and alpha-synuclein accumulation assays

Neuronal lysates were extracted for *in vitro* GCase activity measurements using 4-methylumbelliferyl-β-D-glucopyranoside (Sigma Aldrich, M3633) as described previously ^46^ Alpha-synuclein accumulation was monitored by Western blotting using mouse anti-alpha-synuclein (BD Biosciences, 610787) in whole cell lysates and in Triton-soluble and –insoluble fractions. The fractionation was done as described previously ^47^.

### Standard Protocol Approvals, Registrations, and Patient Consents

The institutional review boards approved the study protocols, and informed consent was obtained from all participants before entering the study. 23andMe participants provided informed consent and participated in the research online, under a protocol approved by the external AAHRPP-accredited IRB, Ethical & Independent Review Services (E&I Review). Cohorts sequenced at McGill university from Columbia University, McGill University (Quebec, Canada and Montpellier, France) and Sheba Medical Center received approval from McGill Institutional Review Board (IRB; A11-M60-21A). The PPMI cohort, AMP-PD initiative cohorts data available for qualified researchers under an agreement and does not require additional internal IRB approval.

### Data availability

All code is available at our git-hub https://github.com/gan-orlab/GALC. Data used in the preparation of this article were obtained from the AMP PD Knowledge Platform (https://www.amp-pd.org) and PPMI (www.ppmi-info.org). All the variants used for the burden analyses are detailed in the provided supplementary tables and the burden analysis can be repeated using these tables. The full GWAS summary statistics for the 23andMe discovery data set will be made available through 23andMe to qualified researchers under an agreement with 23andMe that protects the privacy of the 23andMe participants. Please visit research.23andme.com/collaborate/ for more information and to apply to access the data.

## Results

### Parkinson’s disease risk variants in the *GALC* locus are associated with increased GalCase enzymatic activity

We performed a GWAS to examine the association between common genetic variants and GalCase activity in the Columbia (N=986) and PPMI (N=468) cohorts. Not surprisingly, the strongest signal was in the *GALC* region for both cohorts (Fig. 1A). Applying COJO analysis, signals from two independent *GALC* SNPs were identified in both cohorts (Table 2). The top hits in *GALC* in PPMI and Columbia cohorts were different, however, the top SNP rs429176 in PPMI GalCase GWAS is almost in complete LD (D’=0.99; R^2^=0.97) with the top hit in Columbia cohort rs380142. Moreover, the secondary hits in both cohorts are also in high LD with each other (rs4445832 and rs28533072; D’=0.99). In each cohort, the two independent signals had opposite directions of effect, as one SNP was associated with increased GalCase activity, and the second, independent SNP, was associated with reduced GalCase activity (Table 2). To search for potential secondary hits outside of the *GALC* locus, we performed GWASs for both cohorts adjusted for the top hits in the *GALC* locus. First, we adjusted for the top significant SNP and then repeated the GWAS after adjustment for both independent SNPs in the *GALC* locus (Fig. 1B and C). After adjustment for the top SNP, there were two loci identified in the Columbia University cohort, and two loci in the PPMI cohort that passed Bonferroni correction for multiple comparisons (Supplementary Table 1). However, none of these loci that were found in one of the cohorts was replicated in the other. Similarly, after adjusting for both top SNPs in the *GALC* locus, two loci were associated with GalCase activity in the PPMI cohort but were not replicated in the Columbia University cohort (Supplementary Table 1). In the meta-analysis, only variants in *GALC* locus were associated with GalCase enzymatic activity (Fig. 2A). No other SNPs outside *GALC* locus were associated with GalCase activity in the meta-analysis of the two cohorts before and after adjustments for the top independent hit in the *GALC* locus (Fig. 2B) and for both independent hits (Fig. 2C).

**Table 2.** Conditional and joint analysis identified independent SNPs in GWAS analysis.

| Cohort | SNP | Ref allele | freq | beta | se | p | beta_J | beta_J_se | p_J | LD with PD top hit (rs979812) |
| --- | --- | --- | --- | --- | --- | --- | --- | --- | --- | --- |
| PPMI | rs4445832 | C | 0.4 | 2.33 | 0.09 | $7.12 \times 10^{-91}$ | 1.32 | 0.21 | $1.18 \times 10^{-10}$ | $D' = 0.98 R^2 = 0.69$ |
| PPMI | rs429176 | C | 0.45 | -2.25 | 0.09 | $1.21 \times 10^{-88}$ | -1.32 | 0.2 | $1.73 \times 10^{-11}$ | $D' = 0.63 R^2 = 0.30$ |
| Columbia | rs380142 | C | 0.46 | -1.39 | 0.06 | $4.77 \times 10^{-95}$ | -1.44 | 0.08 | $9.66 \times 10^{-82}$ | $D' = 0.64 R^2 = 0.30$ |
| Columbia | rs28533072 | C | 0.88 | 0.39 | 0.09 | $8.79 \times 10^{-6}$ | 0.57 | 0.09 | $1.68 \times 10^{-10}$ | $D' = 1.0 R^2 = 0.24$ |

GWAS = Genome-wide association studies; Ref Allele = reference allele; freq = minor allele frequency; se- standard error; beta\_J = beta joint analysis, p\_J = p-value joint analysis; SNP = single nucleotide polymorphism.

**Figure 1.**
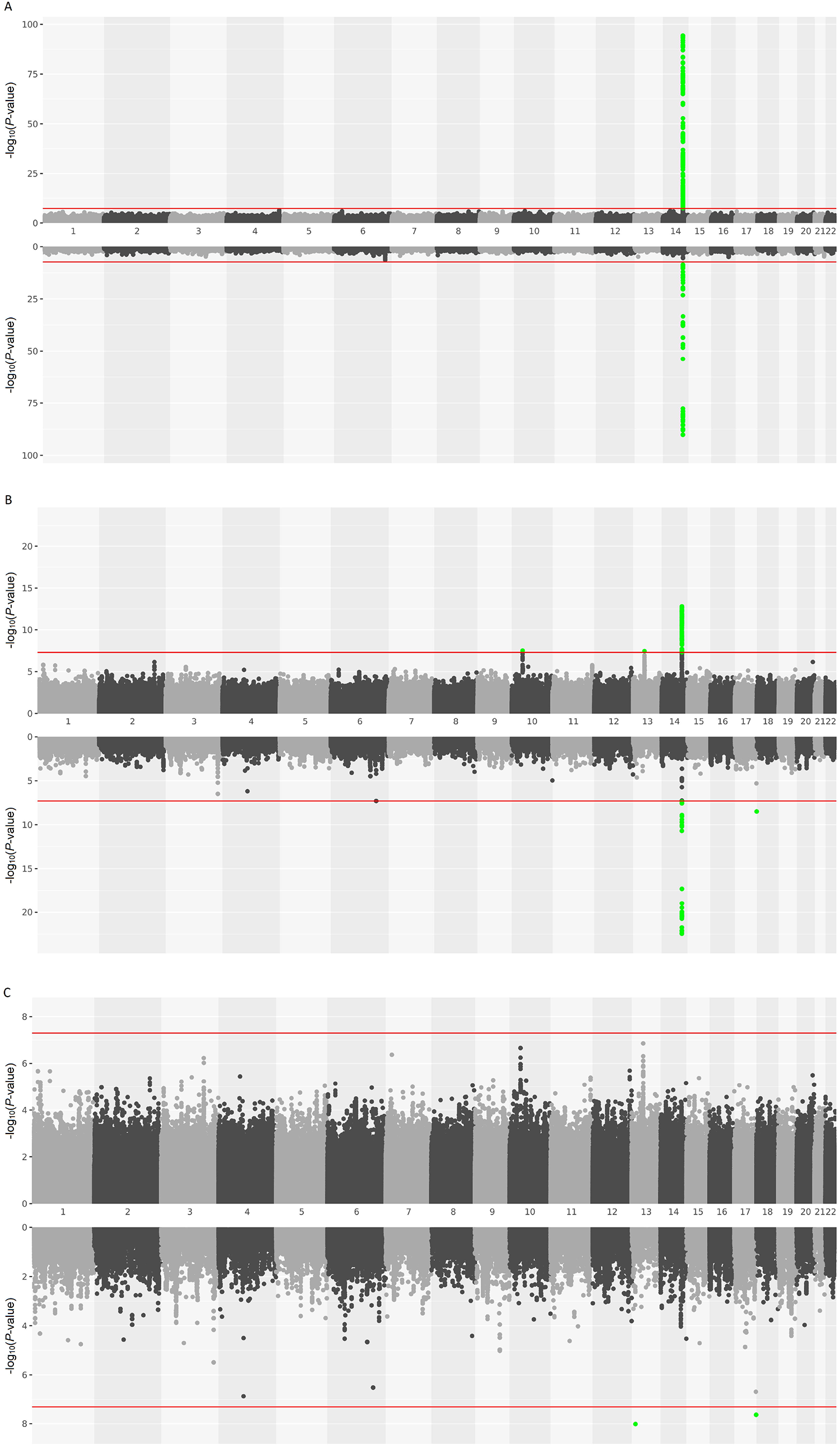
Mirror Manhattan plot of GalCase activity GWAS. The Columbia cohort on the top and the PPMI cohort on the bottom of each plot. The red line indicates GWAS significance threshold. Green dots indicate passing the threshold variants. **A**. GWAS with no adjustments. **B**. GWAS with adjustment for the top SNP associated with GalCase activity. **C**. GWAS with adjustment for the two independent SNPs associated with GalCase activity.

**Figure 2.**
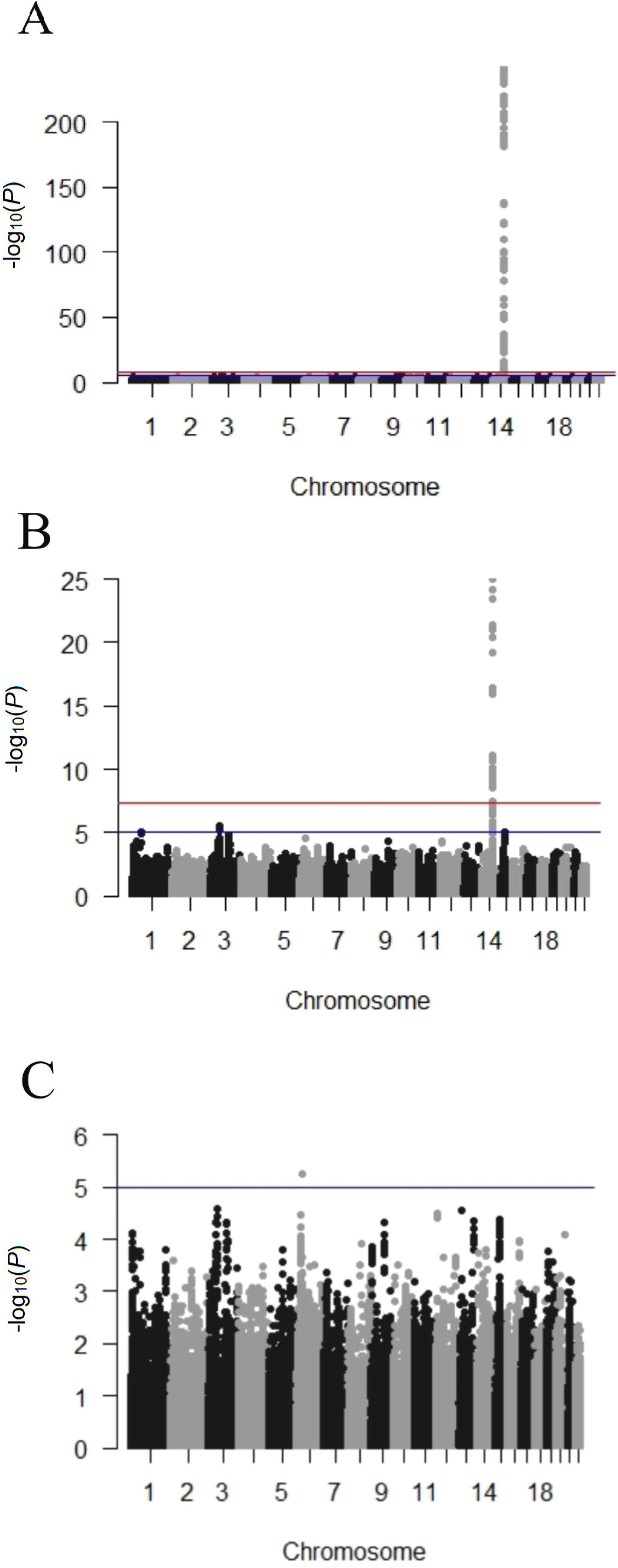
GWAS meta-analysis of GalCase activity between the Columbia and PPMI cohorts. A. Meta-analysis with no adjustments. B. Meta-analysis with adjustment for the top SNP associated with GalCase activity. C. Meta-analysis adjusting for the two independent SNPs associated with GalCase activity.

We then examined the effect of the *GALC* locus SNP rs979812, which was associated with Parkinson’s disease in two previous GWAS meta-analyses ^1,18^, on GalCase activity. This SNP was strongly associated with increased GalCase enzymatic activity (b=1.2; se=0.06; p=5.10E-95). This effect was replicated in both cohorts and in the meta-analysis (Table 3). To exclude a possible effect of PD, we analyzed this SNP in controls only from both cohorts, with similar results (b=1.1; se=0.1; p=2.16E-25). We then examined whether there are coding variants in *GALC* that are associated with GalCase activity. The major allele of the common non-synonymous variant p.I562T (rs398607), which is in partial linkage disequilibrium (D’=0.64) with the top Parkinson’s disease risk variant in this locus, rs979812, was associated with increased *GALC* activity (b=1.65, se=0.05, p=2.70E-242). This variant is associated with Parkinson’s disease but does not reach the level of GWAS significance (b=-0.05, se=0.01, p=1.51E-06). We also found two additional common *GALC* variants which were significantly associated with decreased GalCase activity: p.R184C (b=-1.45, se=0.14, p=4.18e-24) and p.A17T (b=-0.88, se=0.13, p=8.26e-12), but these variants were not associated with Parkinson’s disease. These three common variants were previously reported as modifiers of GalCase enzymatic activity, but they do not cause Krabbe disease in a homozygous state ^48,49^. In conjunction with deleterious *GALC* variants, however, these two variants may cause late-onset Krabbe disease ^49-51^. We also compared GalCase enzymatic activity between cases and controls after merging the PPMI and the Columbia cohorts. We saw slightly increased GalCase activity in cases, which did not reach statistical significance (b=0.03; se=0.029, p=0.29).

**Table 3.**
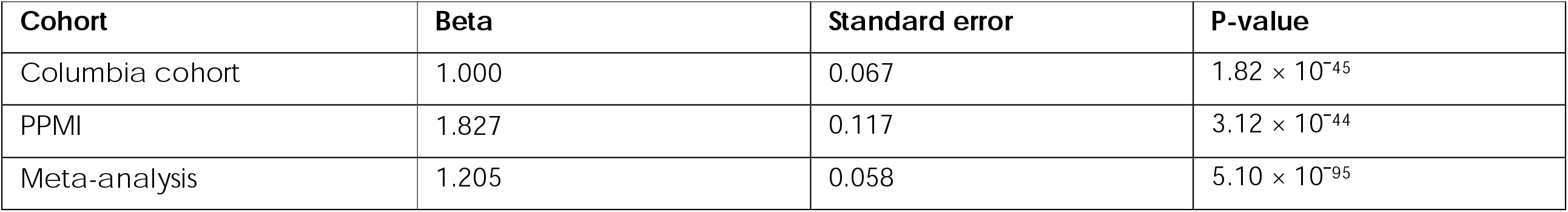
Effe ct of known Parkinson’s disease GWAS locus top hit rs979812 near *GALC* on enzymatic activity.

| Cohort | Beta | Standard error | P-value |
| --- | --- | --- | --- |
| Columbia cohort | 1.000 | 0.067 | $1.82 \times 10^{-45}$ |
| PPMI | 1.827 | 0.117 | $3.12 \times 10^{-44}$ |
| Meta-analysis | 1.205 | 0.058 | $5.10 \times 10^{-95}$ |

### Mendelian randomization supports a potentially causative role of GalCase enzymatic activity in Parkinson’s disease

To examine whether increased GalCase activity may be causal in Parkinson’s disease, we performed MR using the current summary statistics from our GalCase activity analyses as exposure and summary statistics from the most recent Parkinson’s disease GWAS ^1^ as an outcome. We performed MR using the summary statistics from the analysis of the Columbia University cohort alone, the PPMI cohort alone, and their meta-analysis. All three MR analyses demonstrated consistent potentially causative effects on Parkinson’s disease risk (Supplementary Fig. 1, Columbia University: IVW, b=0.029, se=0.011, p=0.006; PPMI: IVW, b=0.022, se=0.006, p=0.0005; meta-analysis: IVW, b=0.025, se=0.007, p-value=0.0008). Taking into account that SNPs in the *GALC* locus are associated with Parkinson’s disease, we applied Steiger filtering to exclude any pleiotropic variants. Moreover, different sensitivity methods did not detect meaningful heterogeneity or pleiotropy (Supplementary Table 2).

### Colocalization suggests that *GALC* association with Parkinson’s disease may be driven by changes in expression

One hypothesis for the association between variants in the *GALC* locus, GalCase activity, and risk of Parkinson’s disease, is that variants in this locus may affect the expression levels of *GALC*, which may lead to increased measured GalCase activity. The top Parkinson’s disease risk-associated SNP, rs979812, was associated with both *GALC* expression and GalCase activity (Fig. 3). Colocalization analysis of the top PD GWAS SNPs with the top GalCase activity GWAS SNPs associated with increased GalCase activity demonstrated colocalization (PPH4>0.8), indicating that increased expression and GalCase activity is associated with Parkinson’s disease. Colocalization analysis with the top GalCase activity GWAS SNPs that were associated with reduced GalCase activity showed no colocalization with PD risk SNPs (PPH4<0.8), suggesting that reduced GalCase expression and activity are not associated with Parkinson’s disease.

**Figure 3.**
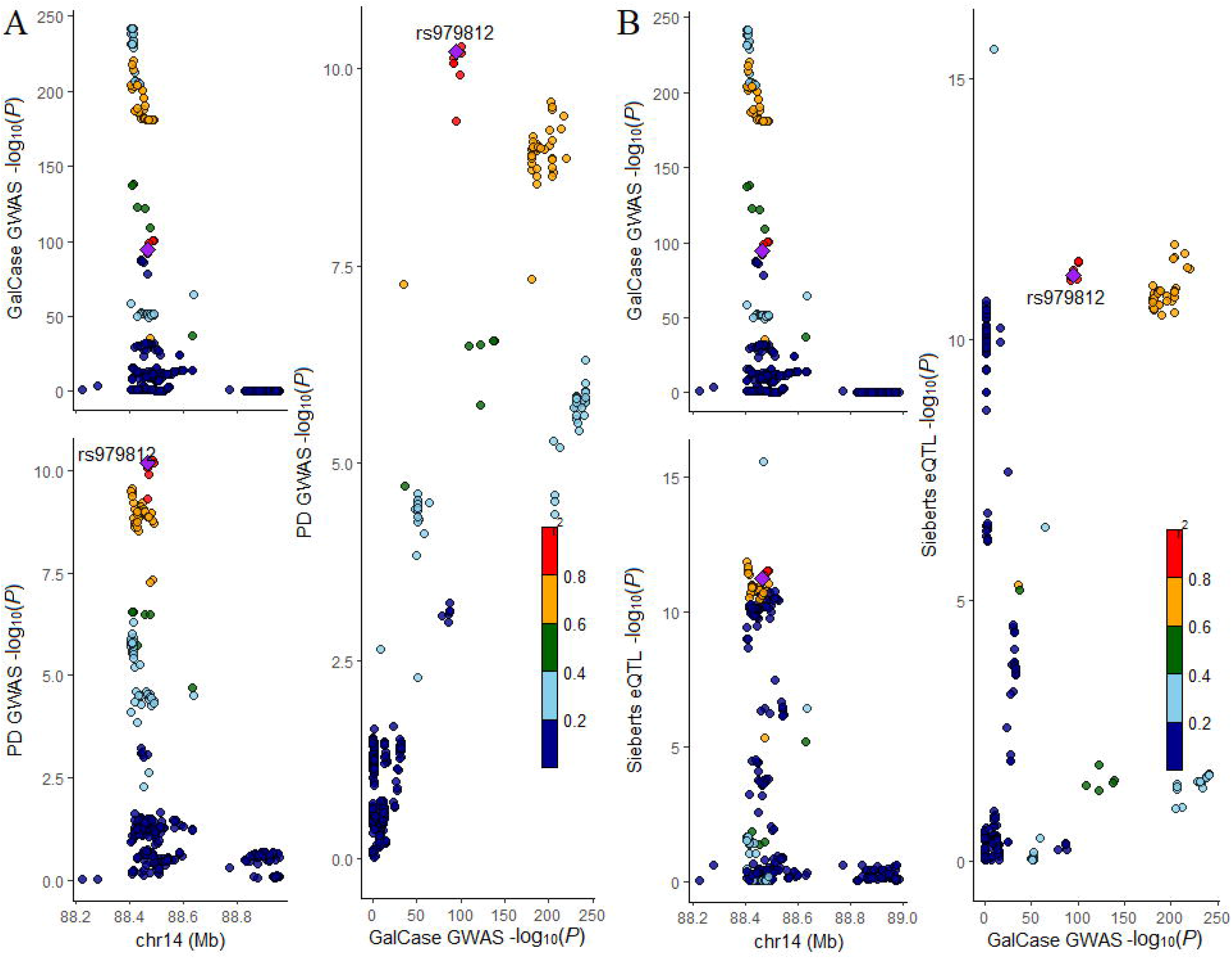
Locus zoom plot comparing *GALC* locus (+/- 500 kb) in Parkinson’s disease GWAS with GalCase GWAS and brain eQTL meta-analysis. A. Parkinson’s disease GWAS plotted together with GalCase activity GWAS meta-analysis. B. Parkinson’s disease GWAS plotted together with brain eQTL meta-analysis.

### No evidence for association of rare *GALC* variants with Parkinson’s disease

We next aimed to examine whether rare *GALC* variants may also be associated with Parkinson’s disease. We performed targeted sequencing in three cohorts and extracted data from a fourth cohort (Table 1) with a total of 5,028 Parkinson’s disease patients and 5,422 controls. In the three cohorts sequenced at McGill University using targeted sequencing, the average coverage of the *GALC* gene was 5539X, with 98% of the nucleotides covered at ≥ 30X. We performed burden analysis using SKAT-O in each of the cohorts separately and then meta-analyzed all cohorts. We did not identify any association between rare *GALC* variants and Parkinson’s disease in any of the cohorts and in the meta-analysis (Supplementary table 3-4).

### Structural analysis of common GalCase variants

The structure of mouse GalCase in complex with substrates and intermediates revealed how the enzyme catalyzes the hydrolysis of galactocerebroside ^52,53^. The protein consists of an unstructured N-terminal 40 a.a. signal peptide, followed by a b-sandwich, TIM barrel, and lectin domains (Fig. 4A). The active site is located in the TIM barrel, adjacent to the lectin domain which binds carbohydrate, thus positioning substrates for cleavage of the glycosidic bond. The structure of human GalCase, predicted using the AlphaFold server ^54^, is highly similar to the experimental mouse GalCase structures (83% sequence similarity). This model allows us to predict the effect of two out of the three missense mutations reported in this study. The p.A17T variant is located in the unstructured signal peptide and therefore its effect cannot be evaluated using our model. On the other hand, the p.R184C and p.I562T variants can be modeled with high certainty. Arg184 is located in the TIM barrel domain and its sidechain is exposed to the solvent (Fig. 4B). The mutation p.R184C causes no steric clash, and would likely not affect substrate binding, given that it is located 33 Å away from the active site. The effect of the p.R184C mutation on GalCase structure is thus likely benign, although we cannot exclude that Arg184 may be involved in some yet unknown protein-protein interaction required for the function or maturation of GalCase. Ile562 is located in the hydrophobic core of the lectin domain (Fig. 4C). The mutation p.I562T would not induce any steric clash, but introduction of a polar amino acid at this position would likely destabilize the fold. Thus, the p.I562T mutation may lead to improper maturation of GalCase, which could interfere with its lysosomal function.

**Figure 4.**
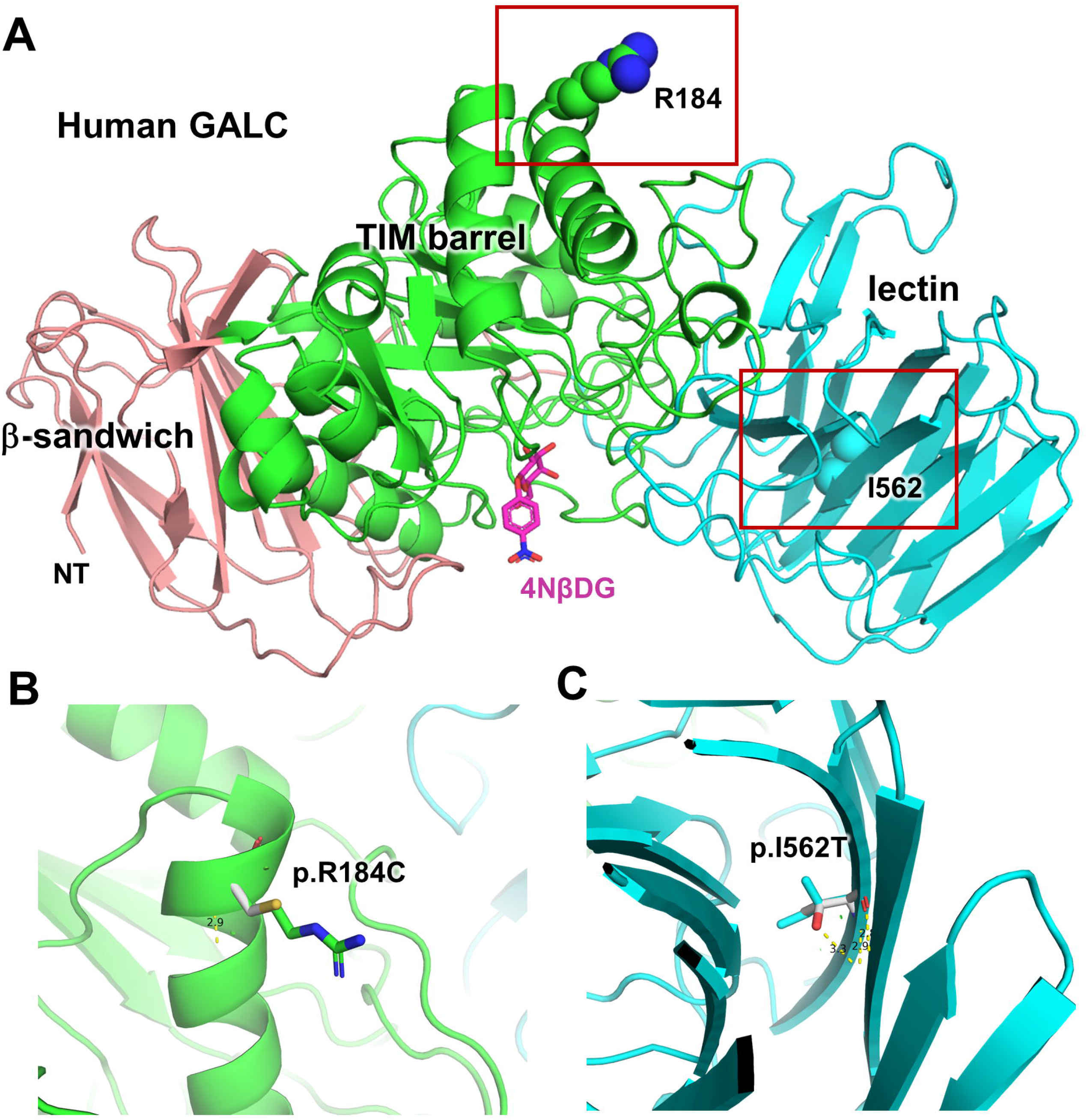
Structural analysis of GALC variants. **(A)** AlphaFold model of human GALC. The structure of mouse GALC bound to the chromogenic substrate 4-nitrophenyl beta-D-galactopyranoside (4NβDG, magenta) was superposed on the human GALC model to indicate the position of the substrate binding site. The side chains of Arg184 and Ile562 are shown as spheres. **(B)** Effect of the mutation p.R184C. The mutated side chains are shown in white. (**C)** Effect of the mutation p.I562T. Predicted hydrogen-bonds are shown in yellow.

### *GALC* knockout does not lead to alterations in GCase activity or to alpha-synuclein accumulation in iPSC-derived Ngn2-induced neurons

Lastly, we sought to examine the effects of altered GalCase activity on Parkinson’s disease-associated phenotypes in iPSC-derived neurons. Considering that several genes involved in interconnected lysosomal lipid metabolism pathways have been linked to Parkinson’s disease in the GWAS studies described above, including *GALC* and *GBA*, we asked whether ablation of the former can affect the activity of the latter. Using an *in vitro* assay, we measured the cleavage of 4-methylumbelliferyl-β-D-glucopyranoside catalyzed by GCase in wild type, *GALC* knockout and *GBA* knockout lysates. As expected, *GBA* knockout lysates showed no GCase activity (Fig. 5A). Wild type and *GALC* knockout lysates had comparable levels of GCase activity (Fig. 5A). Since lysosomes are major sites of alpha-synuclein degradation, we examined the accumulation of monomeric endogenous alpha-synuclein in steady-state lysates of wild type, *GALC* knockout and *GBA* knockout Ngn2-induced neurons and found similar levels (Fig. 5B). Through further analyses of Triton-soluble fractions, we likewise found comparable levels of monomeric alpha-synuclein in all three lines (Supplementary Fig. 2A). Moreover, we observed no high molecular weight alpha-synuclein oligomers in the Triton-insoluble fractions at steady-state (Supplementary Fig. 2B). This observation stands in contrast to a previous study, which reported that *GBA* knockout SH-SY5Y neuroblastoma cells exhibit an accumulation of Triton-insoluble alpha-synuclein oligomers compared to wild type cells ^47^. As controls for these experiments, we analyzed the levels of monomeric and oligomeric alpha-synuclein in dopaminergic neural progenitor cells (DA-NPCs) from an *SNCA* triplication line, as well as CRISPR/Cas9-corrected isogenic control and *SNCA* knockout lines. Consistent with the *SNCA* gene copy numbers in each of these lines, we detected proportionately more Triton-soluble monomeric alpha-synuclein in *SNCA* triplication compared to isogenic control NPC lysates, and none in complete *SNCA* KO lysates (Supplementary Fig. 3A). Even in the *SNCA* triplication line, we found no high molecular weight oligomers at steady state (Supplementary Fig. 3B).

**Figure 5.**
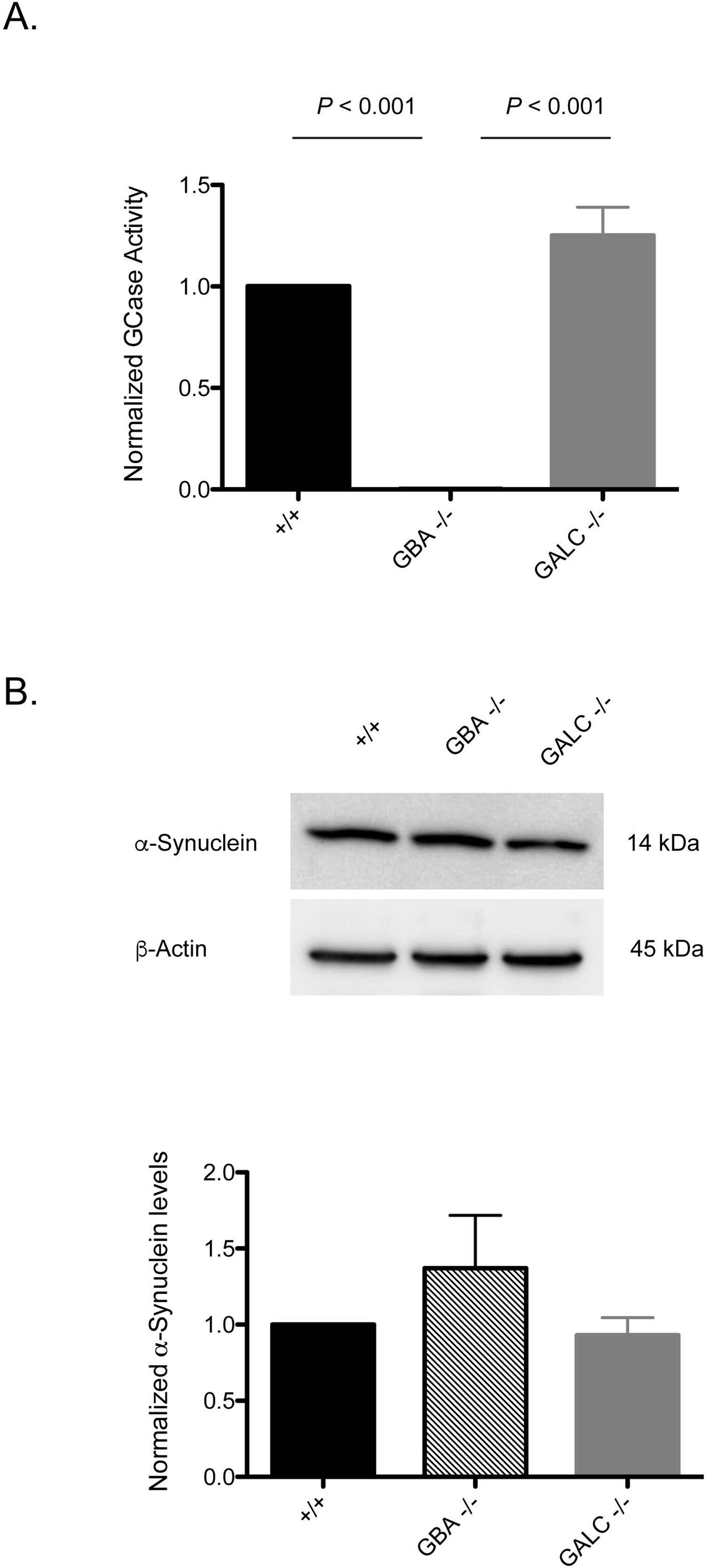
*GALC* loss does not affect GCase activity and monomeric alpha-synuclein accumulation in iPSC-derived Ngn2-induced neurons. (A) GCase activity was measured by 4-methylumbelliferyl- β-D-glucopyranoside cleavage and normalized to the wild-type in three independent replicates and compared with Bonferroni corrected t-tests. There is no statistically significant difference between the +/+ and the *GALC* -/- samples (p > 0.05). (B) Monomeric alpha-synuclein levels were measured in the same lysates. All comparisons are not statistically significant (one-way ANOVA, p = 0.348).

## Discussion

In the current study, we identified two independent signals at the *GALC* locus of variants associated with reduced and increased GalCase activity. We show that the variants associated with increased expression and increased activity of GalCase are also associated with Parkinson’s disease risk. Using MR, we also show that increased GalCase activity may be causal in Parkinson’s disease. Structural analysis of GalCase suggested that the minor allele of p.I562T variant (threonine) may deleteriously affect GalCase function. Since the wildtype amino acid in this allele, isoleucine, is the one associated with Parkinson’s disease risk and with increased GalCase activity, it is possible that this variant is one of the variants driving the association in the *GALC* locus, although there is no full linkage disequilibrium between this variant and the top variant associated with Parkinson’s disease risk. This hypothesis requires additional genetic and functional studies. We did not find an association between rare *GALC* variants and Parkinson’s disease, and KO of GalCase in neuronal models identified no effect on monomeric alpha-synuclein accumulation.

For the vast majority of GWAS loci, the specific gene or genes within each locus that drive the association with Parkinson’s disease are unknown. Our results suggest that in this specific locus, the culprit gene may be *GALC*, specifically through effects on GalCase expression and activity. GalCase works very close and similar to GCase in the lysosomal glycosphingolipid metabolism pathway (Supplementary Fig. 4), which involves other genes and enzymes implicated in Parkinson’s disease, including *SMPD1* ^15^, *ASAH1* ^16^ and *GLA* ^17^. However, the specific mechanism by which these enzymes are affecting the risk of Parkinson’s disease is unclear. Lysosomal genes such as *GBA* work on glycosphingolipids within the lysosomal membrane, and their dysfunction may alter the composition of the lysosomal membrane ^55^. It was hypothesized that these alterations to the membrane composition may affect the ability of the lysosome to internalize and degrade alpha-synuclein ^8^. The involvement of multiple genes from this pathway, including *GALC*, may indicate that maintaining the normal flux within this pathway, and thus maintaining the normal composition of the lysosomal membrane, may be crucial to avoid alpha-synuclein accumulation and the development of Parkinson’s disease. Additional studies are required to test this hypothesis, and in the context of our current results, especially test the effects on increased GalCase activity on the flux in this pathway. Although Parkinson’s disease is associated with decreased activity of GCase, we have demonstrated the opposite effect for GalCase. Another possibility is that trafficking of the enzyme is impaired, leading to reduced lysosomal but increased total activity. Nevertheless, a specific mechanism of how increased GalCase is associated with Parkinson’s disease is yet to be identified. Future studies with variants that increase GalCase expression and activity may be useful for understanding the mechanism behind the association with Parkinson’s disease.

Our Mendelian randomization analysis suggested that increased GalCase activity levels is potentially causal in Parkinson’s disease. These results suggest that reducing GalCase activity may become a target for drug development. When considering this, it is important to remember that three independent studies did not identify a difference in GalCase activity between Parkinson’s disease and controls in the blood ^56,57^ and in brains of Parkinson’s disease patients ^58^. Therefore, if such a strategy of reducing GalCase activity levels will be considered, it should be studied in the context of having *GALC* variants associated with increased activity. Another important point is that Mendelian randomization has some limitations. It is dependent on the quality of the GWASs used for the summary statistics, and despite the tools used to exclude pleiotropic variants, there could still be residual, cryptic pleiotropy ^59^. Therefore, our results should be confirmed in additional, independent studies, whether population studies or functional studies in relevant models. Knockout of *GALC* had no effect on the monomeric alpha-synuclein accumulation in iPSC-derived Ngn2-induced neurons. These results are in line with our results showing that the variants p.R184C and p.A17T that may cause adult-onset Krabbe disease when inherited together with other deleterious *GALC* variants, thus reducing GalCase activity, are not associated with Parkinson’s risk. In addition, our analyses of rare *GALC* variants, as well as previous studies ^60^, suggest minor or no role of rare *GALC* variants in Parkinson’s disease development.

Krabbe disease is a lysosomal storage disorder caused by biallelic deleterious variants in *GALC*, typically manifesting in infancy with death before the age of 2 years, but also with more benign late-onset forms ^61,62^. Several variants in *GALC* are leading to pseudodeficiency, carriers of these variants have deficient GalCase which do not lead to Krabbe disease ^63^. One of these variants, p.I562T, is considered as a benign variant for Krabbe disease, yet the threonine residue, although not pathogenic on its own, may be a modifier of Krabbe disease severity ^49^. This also corresponds well with our findings, showing that the activity of the major allele with the amino acid isoleucine is increased compared to the minor allele threonine. The isoleucine allele was also associated with Parkinson’s disease risk, although this association is below GWAS-corrected statistical significance threshold. Whether this allele is driving the association in the *GALC* region is still unclear, and it is possible that other, non-coding variants in this region, drive the effects on GalCase activity and risk of Parkinson’s disease. Comprehensive genomic and direct functional assessment of variants located in the promoter or enhancers of GalCase will be required to identify the specific variant or variants that drive the association and the relevant mechanism.

Our study has several limitations. In some of our cohorts, there was a significant difference in sex between Parkinson’s disease patients and controls. This limitation was addressed by adjustment in the regression model with sex as a covariate, as well as other covariates. We have also adjusted for ethnicity in the analysis of the Columbia cohort since there were people with European and Ashkenazi Jewish ancestry. The restriction of our analysis for individuals mainly of European and Ashkenazi Jewish ancestry is also a limitation. In this study, GalCase activity was measured from DBS, including in the PPMI cohort where they were prepared from frozen blood. To account for this difference in preparation, we adjusted for white blood cell count. However, since DBS does not measure the enzymatic activity within the lysosomal environment in live cells, future studies will be required to replicate our results in different models using different methods for measuring enzymatic activity. In the current study we have measured GalCase activity in DBS from peripheral blood, which might not perfectly represent GalCase activity in the brain. However, based on GCase studies, measured using the same method, we know that GCase activity measured in DBS is associated with *GBA* genotype and the results were reproducible across different studies ^14,56^ and with longitudinal measurements ^21^. Moreover, previous findings on GCase activity in brain ^64^ were consistent with the DBS results taken from peripheral blood ^14^. Taking into account these data, it is plausible that the activity of these lysosomal enzymes in blood is a valuable and reproducible proxy for their activity in the brain. Nevertheless, there are currently no studies that compared activity of lysosomal enzymes simultaneously in brain tissue and peripheral blood. The Mendelian randomization analysis that we performed also has several limitations, some of which we described above. The PPMI cohort was included in the Parkinson’s disease meta-analysis which we used as outcome. This will have a very minor effect or no effect at all since the PPMI cohort represents a very small fraction of the Parkinson’s GWAS meta-analysis. In addition, the GWAS on GalCase activity was performed in Parkinson’s disease cases and healthy controls. Even though we account for it by adjusting for disease status in the regression model, it could still create some biases. Another limitation related to GalCase is that we present results on activity measured from blood, yet GalCase activity could behave differently in brain tissues.

Another limitation of our study is that the cell model we used study is a loss-of-function model, whereas our results indicate that a gain-of-function, i.e., increased expression and activity of GalCase, are associated with risk of PD. In addition, our models do not examine the role of specific variants we identified in this study and the mechanisms by which they potentially influence GalCase expression, activity and risk of PD. Therefore, future studies with additional models are necessary to further delineate the potential mechanisms underlying the association between GalCase expression and activity and risk of PD. Such studies could include the following: a) Overexpression models, preferably in PD-relevant cell models such as neurons and microglia. This can be achieved, for example, by using CRISPR-cas-based transcriptional activators in induced pluripotent stem cells (iPSCs), followed by reprogramming to neuron and microglia cells. Using such GalCase overexpression models, different assays could then be performed, including examining alpha-synuclein accumulation, uptake, degradation and phosphorylation, with and without adding alpha-synuclein pre-formed fibrils (PFFs), effects on GCase activity using lysosomal specific substrates, and general effects on lysosomal structure and function. b) models with specific genetic variants implicated in the current study, whether non-coding variants that affect GalCase expression and activity, and/or coding variants that might affect the structure and function of GalCase including p.A17T, p.R184C and p.I562T. Since these variants are not rare, they can be modeled from patient-derived iPSCs and corrected with CRISPR-cas9 to get isogenic controls. Such models will isolate the effects of these variants in these cell lines, and then using similar assays mentioned above to study their effects on PD-related mechanisms. Other experiments in these models, including localization of GalCase in the lysosome, direct measurements of GalCase activity and others, can also shed more light on the potential mechanisms that may link these variants to PD.

To conclude, our findings support a role for the *GALC* gene in risk of Parkinson’s disease, possibly through alterations in GalCase activity due to genetic variants. These findings suggest that reducing GalCase activity could be considered for pre-clinical translational studies in Parkinson’s disease, yet it will likely be relevant only for subgroups of patients with specific genetic background and GalCase activity profiles. Due to the limitations mentioned, and before embarking on translational studies, further genetic and functional studies are required to replicate our findings.

## Abbreviations

AMP-PD: Accelerating Medicines Partnership – Parkinson Disease
CADD: Combined Annotation Dependent Depletion
COJO: Conditional and joint analyses
DBS: dried blood spotsuantitative trait loci
GalCase: galactosylceramidase
GalCer: galactosylceramide
GCase: glucocerebrosidase
GWAS: genome-wide association study
HBS: Harvard Biomarkers Study
IVW: inverse-variance weighted
LBD: Lewy body dementia
LC-MS/MS: liquid chromatography-tandem mass spectrometry
MAF: minor allele frequency
MIPs: Molecular Inversion Probes
MR: Mendelian randomization
MR-PRESSO: MR pleiotropy residual sum and outlier
mQT: methylation quantitative trait loci
NIA: National Institute on Aging
PDBP: Parkinson’s disease Biomarkers Program
PPMI: Parkinson’s Progression Markers Initiative
SKAT-O: sequence kernel association optimal unified test
STEADY-PD3: Study of Isradipine as a Disease Modifying Agent in Subjects With Early Parkinson Disease, Phase 3
sQTL: splicing quantitative trait loci

## Acknowledgements

We would like to thank the participants in the different cohorts for contributing to this study. Data used in the preparation of this article were obtained from the AMP PD Knowledge Platform. For up-to-date information on the study, visit https://www.amp-pd.org. AMP PD – a public-private partnership – is managed by the FNIH and funded by Celgene, GSK, the Michael J. Fox Foundation for Parkinson’s Research, the National Institute of Neurological Disorders and Stroke, Pfizer, Sanofi, and Verily. Genetic data used in preparation of this article were obtained from the Fox Investigation for New Discovery of Biomarkers (BioFIND), the Harvard Biomarker Study (HBS), the Parkinson’s Progression Markers Initiative (PPMI), the Parkinson’s Disease Biomarkers Program (PDBP), the International LBD Genomics Consortium (iLBDGC), and the STEADY-PD III Investigators. BioFIND is sponsored by The Michael J. Fox Foundation for Parkinson’s Research (MJFF) with support from the National Institute for Neurological Disorders and Stroke (NINDS). The BioFIND Investigators have not participated in reviewing the data analysis or content of the manuscript. For up-to-date information on the study, visit michaeljfox.org/news/biofind. The HBS is a collaboration of HBS investigators [full list of HBS investigators found at https://www.bwhparkinsoncenter.org/biobank/ and funded through philanthropy and NIH and Non-NIH funding sources. The HBS Investigators have not participated in reviewing the data analysis or content of the manuscript. PPMI – a public-private partnership – is funded by the Michael J. Fox Foundation for Parkinson’s Research and funding partners, including [list the full names of all of the PPMI funding partners found at www.ppmi-info.org/fundingpartners]. The PPMI Investigators have not participated in reviewing the data analysis or content of the manuscript. For up-to-date information on the study, visit www.ppmi-info.org. PDBP consortium is supported by the NINDS at the National Institutes of Health. A full list of PDBP investigators can be found at https://pdbp.ninds.nih.gov/policy. The PDBP investigators have not participated in reviewing the data analysis or content of the manuscript. Genome Sequencing in Lewy Body Dementia and Neurologically Healthy Controls: A Resource for the Research Community.” was generated by the iLBDGC, under the co-directorship by Dr. Bryan J. Traynor and Dr. Sonja W. Scholz from the Intramural Research Program of the U.S. National Institutes of Health. The iLBDGC Investigators have not participated in reviewing the data analysis or content of the manuscript. For a complete list of contributors, please see: bioRxiv 2020.07.06.185066; doi: https://doi.org/10.1101/2020.07.06.185066. STEADY[PD III is a 36[month, Phase 3, parallel group, placebo[controlled study of the efficacy of isradipine 10 mg daily in 336 participants with early Parkinson’s Disease that was funded by the NINDS and supported by The Michael J Fox Foundation for Parkinson’s Research and the Parkinson’s Study Group. The STEADY-PD III Investigators have not participated in reviewing the data analysis or content of the manuscript. The full list of STEADY PD III investigators can be found at: https://clinicaltrials.gov/ct2/show/NCT02168842. Some data was also obtained using PPMI database (www.ppmiinfo.org/data). For up-to-date information on the study, visit www.ppmiinfo.org.” “PPMI – a public-private partnership – is funded by the Michael J. Fox Foundation for Parkinson’s Research and funding partners, including (list the full names of all of the PPMI funding partners found at www.ppmiinfo.org/fundingpartners). We would also like to thank the research participants and employees of 23andMe for making this work possible. The full GWAS summary statistics for the 23andMe discovery data set will be made available through 23andMe to qualified researchers under an agreement with 23andMe that protects the privacy of the 23andMe participants. Please visit research.23andme.com/collaborate/ for more information and to apply to access the data. The results published here are in whole or in part based on data obtained from the AD Knowledge Portal (https://adknowledgeportal.org). Study data were provided by the Rush Alzheimer’s Disease Center, Rush University Medical Center, Chicago, where data collection was supported through funding by NIA grants P30AG10161, R01AG15819, R01AG17917, R01AG36836, R01AG48015, U01AG46152, the Illinois Department of Public Health (ROSMAP), and the Translational Genomics Research Institute (genomic). The Mayo Clinic Alzheimers Disease Genetic Studies, led by Dr. Nilufer Taner and Dr. Steven G. Younkin, Mayo Clinic, Jacksonville, FL where data collection was supported through funding by NIA grants P50 AG016574, R01 AG032990, U01 AG046139, R01 AG018023, U01 AG006576, U01 AG006786, R01 AG025711, R01 AG017216, R01 AG003949, NINDS grant R01 NS080820, CurePSP Foundation, and support from Mayo Foundation. Study data includes samples collected through the Sun Health Research Institute Brain and Body Donation Program of Sun City, Arizona. The Brain and Body Donation Program is supported by the National Institute of Neurological Disorders and Stroke (U24 NS072026 National Brain and Tissue Resource for Parkinsons Disease and Related Disorders), the National Institute on Aging (P30 AG19610 Arizona Alzheimers Disease Core Center), the Arizona Department of Health Services (contract 211002, Arizona Alzheimers Research Center), the Arizona Biomedical Research Commission (contracts 4001, 0011, 05-901 and 1001 to the Arizona Parkinson’s Disease Consortium) and the Michael J. Fox Foundation for Parkinsons Research. And, the CommonMind Consortium supported by funding from Takeda Pharmaceuticals Company Limited, F. Hoffmann-La Roche Ltd and NIH grants R01MH085542, R01MH093725, P50MH066392, P50MH080405, R01MH097276, RO1-MH-075916, P50M096891, P50MH084053S1, R37MH057881, AG02219, AG05138, MH06692, R01MH110921, R01MH109677, R01MH109897, U01MH103392, and contract HHSN271201300031C through IRP NIMH. Brain tissue for the study was obtained from the following brain bank collections: the Mount Sinai NIH Brain and Tissue Repository, the University of Pennsylvania Alzheimer’s Disease Core Center, the University of Pittsburgh NeuroBioBank and Brain and Tissue Repositories, and the NIMH Human Brain Collection Core. CMC Leadership: Panos Roussos, Joseph Buxbaum, Andrew Chess, Schahram Akbarian, Vahram Haroutunian (Icahn School of Medicine at Mount Sinai), Bernie Devlin, David Lewis (University of Pittsburgh), Raquel Gur, Chang-Gyu Hahn (University of Pennsylvania), Enrico Domenici (University of Trento), Mette A. Peters, Solveig Sieberts (Sage Bionetworks), Thomas Lehner, Stefano Marenco, Barbara K. Lipska (NIMH). This work was supported in part by the Intramural Research Programs of the National Institute on Aging (NIA). ZGO is supported by the Fonds de recherche du Québec - Santé (FRQS) Chercheurs-boursiers award, in collaboration with Parkinson Quebec, and is a William Dawson Scholar. The access to part of the participants for this research has been made possible thanks to the Quebec Parkinson’s Network (http://rpq-qpn.ca/en/). KS is supported by a post-doctoral fellowship from the Canada First Research Excellence Fund (CFREF), awarded to McGill University for the Healthy Brains for Healthy Lives initiative (HBHL) and FRQC post-doctoral fellowship.

## Supplementary material

Supplementary Table 1. Loci significantly associated with GALC activity in Columbia and PPMI cohorts outside of GALC locus

Supplementary Table 2. Mendelian randomization between GALC and Parkinson’s disease

Supplementary Table 3. SKAT-O analysis of rare (MAF < 1 %) *GALC* variants

Supplementary Table 4. All rare (MAF<1%) *GALC* variants in 4 cohorts

Supplementary Figure 1. MR analysis between exposure (GalCase activity) and outcome (Parkinson’s disease risk).

Supplementary Figure 2. *GALC* loss does not alter Triton-soluble and -insoluble alpha-synuclein levels in iPSC-derived Ngn2-induced neurons.

Supplementary Figure 3. Analysis of Triton-soluble and insoluble alpha-synuclein levels in NPCs with different *SNCA* gene copy numbers.

Supplementary Figure 4. GCase and GalCase role in sphingolipid metabolism. Supplementary material is available at Brain online

