## Supplementary Fig. for "*GALC* variants affect galactosylceramidase enzymatic activity and risk of Parkinson’s disease"

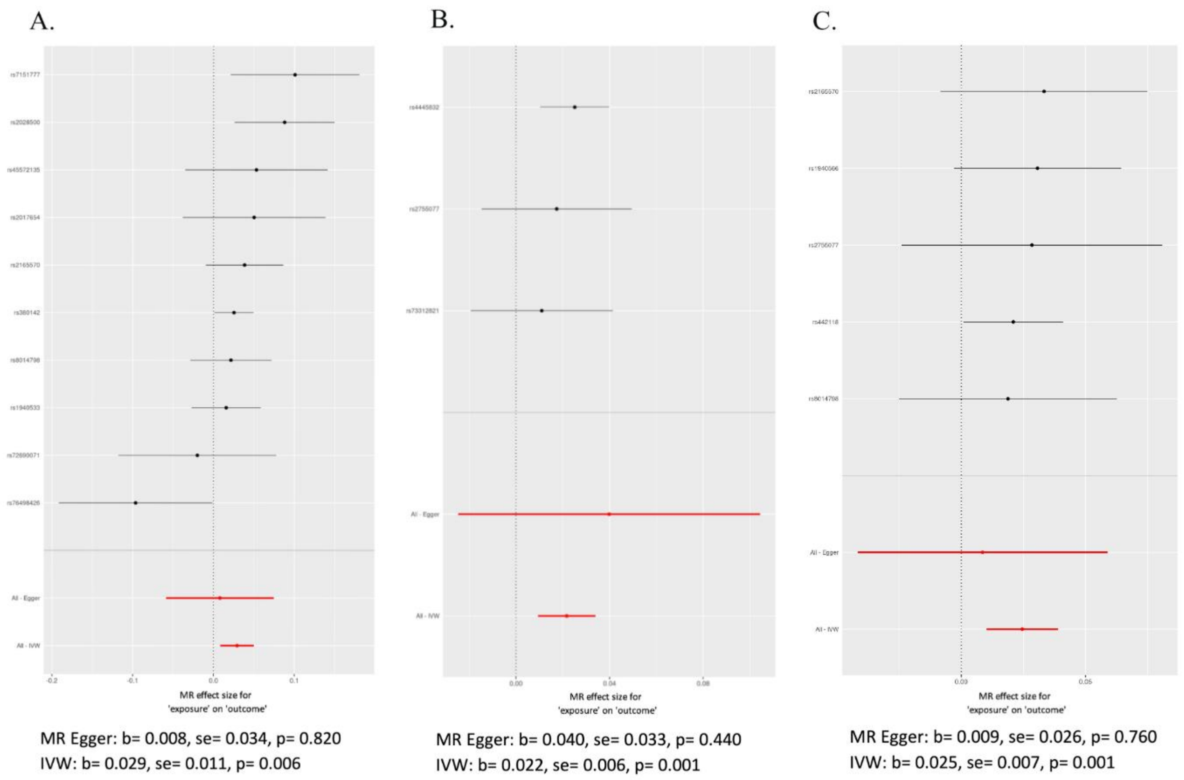


**Supplementary Figure 1. MR analysis between exposure (GalCase activity) and outcome (Parkinson's disease risk).**

A. GalCase activity GWAS in the Columbia cohort as exposure; B. GalCase activity GWAS in PPMI cohort as exposure. C. GalCase activity GWAS meta-analysis as exposure. Abbreviations: Mendelian randomization-Egger – MR-Egger; Inverse variance weighted – IVW; b – beta; se – standard error.

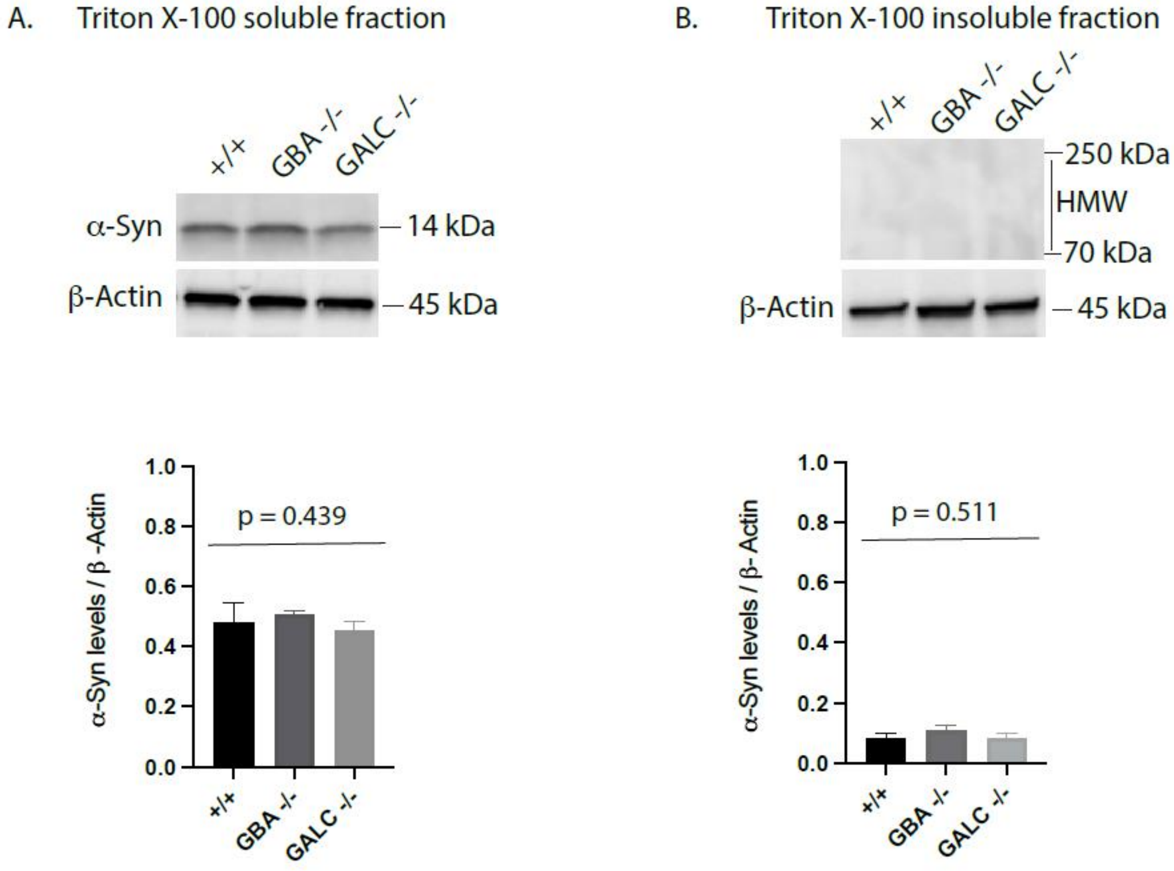


**Supplementary Figure 2. *GALC* loss does not alter Triton-soluble and -insoluble alpha--synuclein levels in iPSC-derived Ngn2-induced neurons.**

(A) Analysis of Triton-soluble monomeric alpha-synuclein in +/+, *GBA* -/- and *GALC* -/- Ngn2 neurons (one-way ANOVA, p= 0.439). (B) Analysis of high molecular weight Triton-insoluble oligomeric alpha-synuclein in +/+, *GBA* -/- and *GALC* -/- Ngn2 neurons (one-way ANOVA, p= 0.511).

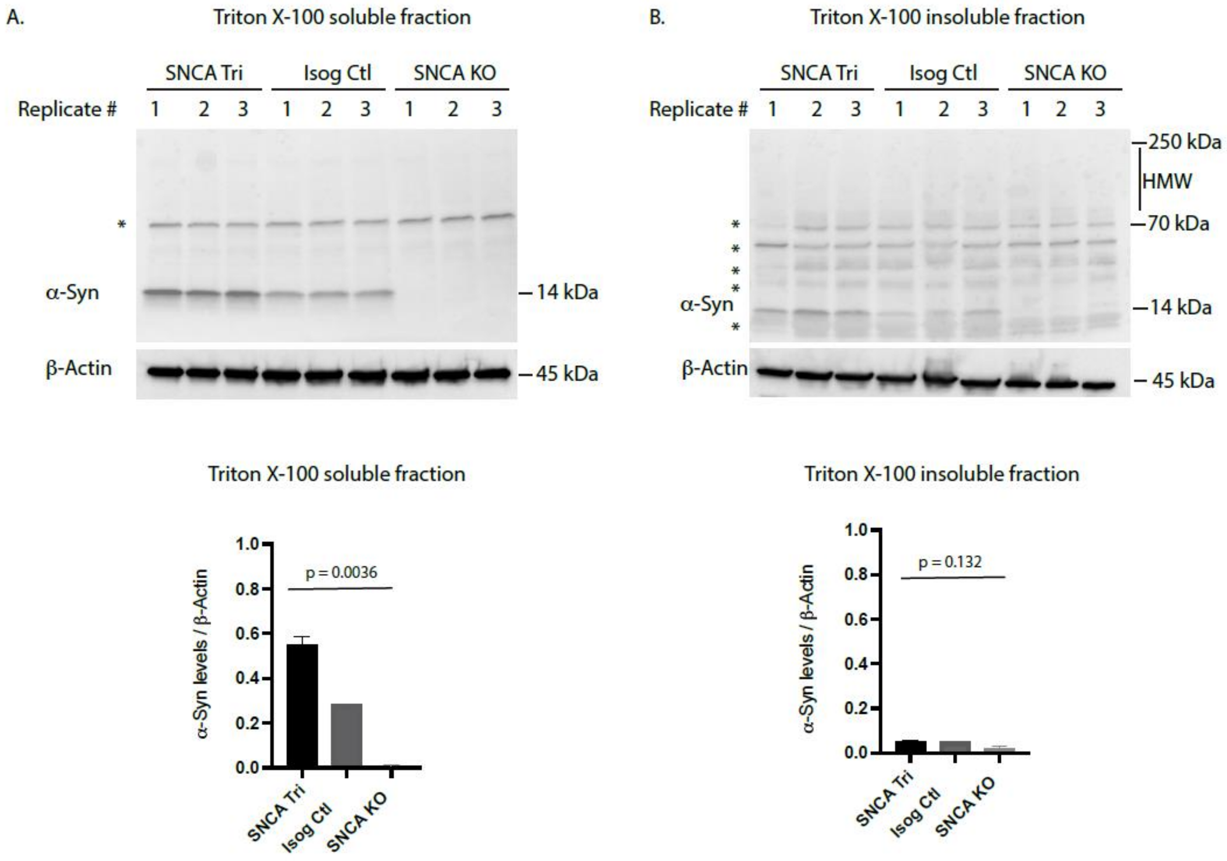


**Supplementary Figure 3. Analysis of Triton-soluble and insoluble alpha-synuclein levels in NPCs with different *SNCA* gene copy numbers.**

(A) Analysis of Triton-soluble monomeric alpha-synuclein in *SNCA* triplication, isogenic control and *SNCA* KO dopaminergic NPCs (one-way ANOVA, p= 0.0036). (B) Analysis of high molecular weight Triton-insoluble oligomeric alpha-synuclein in *SNCA* triplication, isogenic control and *SNCA* KO dopaminergic NPCs (one-way ANOVA, p= 0.132).

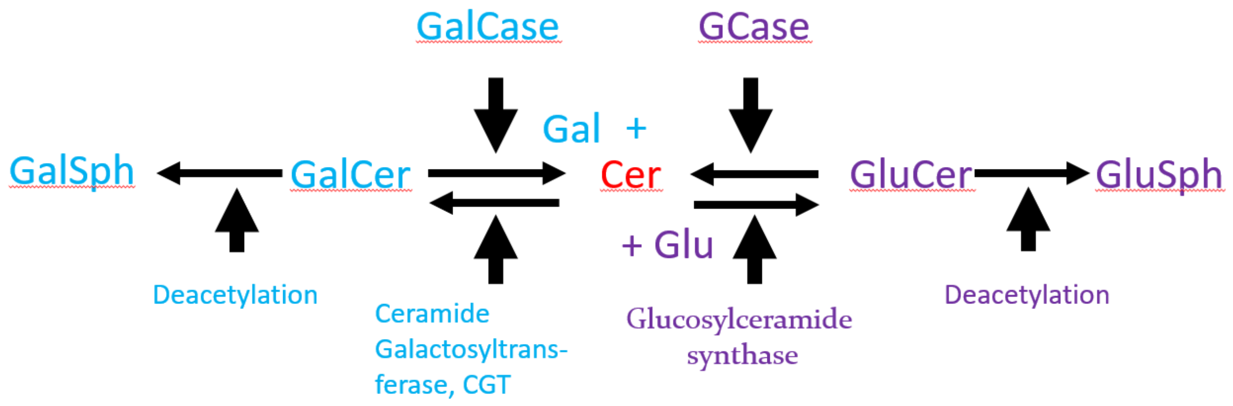


**Supplementary Figure 4. GCase and GalCase role in sphingolipid metabolism.**

The figure depicts a part of the lysosomal ceramide metabolism pathway. *GBA* encodes GCase and *GBA* variants are the most common genetic risk factor of Parkinson's disease. GalCase works similarly to GCase in the lysosomal glycosphingolipid metabolism pathway. Therefore, plausibly plays role in Parkinson's disease pathogenesis. Gal – galactose; Cer – ceramide; Glu – glucose; GalCase – galactosylceramidase; GalCer – galactosylceramide; GCase – glucocerebrosidase; GluCer – glucosylceramide; GluSph – glucosylsphingosine; GalSph – galactosylsphingosine
